# Cell cycle abnormality is a cellular phenotype in OCD

**DOI:** 10.1101/2020.03.31.20041368

**Authors:** Pravallika Manjappa, Srinivas Balachander, Safoora Naaz, Ravi Kumar Nadella, Tulika Shukla, Pradip Paul, Meera Purushottam, YC Janardhan Reddy, Sanjeev Jain, Biju Viswanath, Reeteka Sud

## Abstract

Abnormal indices of cell cycle regulation have been reported in multiple psychiatric disorders. Though reports specific to obsessive compulsive disorder (OCD) are scant, numerous studies have highlighted partly common underlying biology in psychiatric disorders, cell cycle regulation being one such process. In this study, we therefore aimed to explore cell cycle in OCD. To the best of our knowledge, this is the first study to investigate these effects in OCD. We also evaluated the effect of *in vitro* fluoxetine, commonly used serotonin reuptake inhibitor (SRI) in OCD patients, on cell cycle regulation. The effects of both disease (OCD) and treatment (SRI) were assessed using lymphoblastoid cell lines (LCLs), derived from OCD patients and healthy controls, as a model system. LCLs were treated with 10μM of fluoxetine for 24 hours, and the percentage of cells in each phase of the cell cycle was determined by flow cytometry. We observed a lower proportion of cells in the G2/M phase in OCD cases than controls. The findings suggest that cell cycle dysregulation could be peripheral cellular phenotype for OCD. Among cases, all of whom had been systematically characterized for SRI treatment response, LCLs from non-responders to SRI treatment had a lower proportion of cells in G2/M phase than responders.

## 1. Introduction

Characterized by obsessions (intrusive, unwanted thoughts) and compulsions (performing highly ritualized behaviours intended to neutralize negative thoughts and emotions resulting from the obsessions), Obsessive Compulsive Disorder (OCD) is a common psychiatric disorder affecting almost 2% of the general population. Selective serotonin reuptake inhibitors (SRIs) form the mainstay of treatment, but many patients fail to gain therapeutic benefit (Goddard, Shekhar, Whiteman, & Mcdougle, 2008). Clearly, cellular parameters that can serve as biomarkers for OCD, associated with the disease as well as treatment response, are critically needed.

Fluoxetine is a commonly used SRI in the treatment of OCD and influences many cellular parameters, including cell proliferation (Serafeim et al., 2003; Volpe, Ellison, Parchment, Grieshaber, & Faustino, 2003). A recent study reported an increase in the proportion of G0/G1 mouse hippocampal neurons, along with a reduction of G2/M and S phase cells at 48 hrs and 96 hrs of treatment with fluoxetine (Solek et al., 2019). SRIs act through serotonin and dopamine transporters (Lima & Urbina, 2002; Sanders-bush, Hazelwood, 2003), and influence serotonin receptors (Gladkevich, Kauffman, & Korf, 2004); both of which are expressed on lymphocytes. Our lab, and others, have used lymphoblastoid cell lines (LCLs) (derived from lymphocytes) to study cell cycle regulation and cell proliferation, and the effect of SRI response in neuropsychiatric disorders (Breitfeld et al., 2016; Breitfeld, Scholl, Steffens, Laje, & Stingl, 2017; Ashok et al., 2019; Paul et al., 2020).

Abnormal indices of cell cycle regulation have been reported in several psychiatric disorders. Though reports specific to OCD are scant, genomic findings have highlighted partly common underlying biology in psychiatric disorders (Vaidya et al. 2007; Wang et al. 2010; Ashok et al., 2019; Paul et al., 2020). Therefore, we examined the cell cycle in lymphocytes of patients with OCD and compared them with those of healthy controls. In addition, we wanted to examine if fluoxetine *in vitro* has a differential effect on cell cycle in OCD lymphocytes, based on clinical treatment response.

## 2. Materials and Methods

### 2.1 Subjects & assessment of clinical treatment response

All patients with OCD (N=11) were recruited from the Speciality OCD clinic of the National Institute of Mental Health and Neuro Sciences (NIMHANS, Bangalore). The Mini-International Neuropsychiatric Interview (MINI) and Yale-Brown Obsessive-Compulsive Scale (Y-BOCS) were used to diagnose and assess the patients clinically. Individuals (N=8) with no personal and family history of psychiatric illnesses were age-matched with OCD patients and recruited as controls for the study. The study was approved by the Ethics Committee of NIMHANS. Prior to participation, informed written consent was taken. Among the patients, 5 had responded clinically to SRIs, whereas 3 did not. Three patients were partial responders and were excluded from the analysis of treatment response. Responders were those who had >35% reduction in YBOCS (and CGI-I 1 or 2) following an SRI trial; non-responders were those with <25% reduction (and CGI-I >3) even after at least 2 SRI trials (Mataix-Cols et al., 2016; Shukla et al., 2020). All patients were on treatment with SRIs at the time of sample collection.

### 2.2 Generation and characterisation of LCLs

Venous blood samples of patients and controls were collected to isolate peripheral blood mononuclear cells (PBMCs) using SepMate™ kit (Stem Cell Technologies), using the protocol adapted from Ashok et al., 2019. LCLs were generated from PBMCs by transformation using Epstein-Barr virus (Jung, 2013). All cell lines were cultured in RPMI-1640 (HiMedia) medium containing 20% heat-inactivated fetal bovine serum (GIBCO) and 1% Penicillin-Streptomycin (GIBCO); as suspension cultures in a humidified environment at 37°C and 5% CO_2_. The cell lines used had a doubling time of 48-72 hours, and those between passages 3 and 10 were used for the study.

### 2.3 Fluoxetine treatment and cell cycle analysis

LCLs were plated at a density of 10^6^ cells/ml and treated *in vitro* with vehicle or 10μM Fluoxetine (Sigma) for 24 hours after which they were pelleted, washed with 1X phosphate buffer saline (PBS), fixed with 70% ethanol and stored at 4° C for a minimum of 24 hours. Fixed cells were washed and incubated for 30 min at 37° C in 1X PBS solution containing RNase (Invitrogen - 40μg/ml) and Propidium Iodide (Invitrogen - 15μg/ml). The 24-hour fluoxetine treatment protocol was adapted from Porton et al., 2013. Immunophenotyping of LCLs was performed, which showed that they were positive for CD19 (B cell marker) and negative for CD3 (T cell marker) and CD56 (NK cell marker). The experiment was performed in triplicates for each cell line.

Cell cycle analysis was performed by flow cytometry to estimate the proportion of cells in different phases of the cell cycle – G0/G1, S, and G2/M. The percentage of cells in each cell cycle phase was determined using the software FloJo (V10). The scatter plot was gated to include only singlet cells for analysis.

### 2.4 Statistical analysis

Statistical analysis was performed using GraphPad Prism (Version 8.2.1) and R (base packages, version 3.6.2). A 2×2 mixed model ANOVA with post-hoc pairwise comparisons (Tukey’s HSD) were used to compare the different conditions, groups & their interaction. All values <0.05 were considered significant.

## 3. Results

### 3.1 G2/M phase of cell cycle showed differences in control vs OCD cases

Cell cycle analysis in LCLs (vehicle- and fluoxetine-treated) from 8 control and 11 OCD patients was performed and analysed using flow cytometry. Distribution of cells in G0/G1, S and G2/M phases of the cell cycle was determined by propidium iodide staining.

As shown in Table 1, the main effects of “group” and “treatment with fluoxetine” were significant predictors for the percentage of cells, but only for the G2/M phase. The estimated marginal means (EMM) with 95% confidence intervals for cases vs controls was 11.8 (10.5-13.1) vs 14.1 (12.8-15.3), F(1,17)=6.88 & p=0.017; and for vehicle vs fluoxetine-treated lines was 14.1 (12.8-15.3) vs 11.7(10.5-13.0), F(1.17)= 5.97 & p=0.026. Their interaction (Group * Treatment) was not significant, indicating that the effect of fluoxetine treatment was the same in LCLs from OCD cases and healthy controls. These findings are similar to other studies (Lin et al., 2016; Solek et al., 2019). There were no other significant differences in the post hoc comparisons performed using Tukey’s HSD method (Table 2).

**Table 1:** Results of 2×2 Mixed ANOVA comparing cell lines from cases (n=11) vs controls (n=8), with and without treatment with fluoxetine.

|  | G0/G1 Phase |  | S Phase |  | G2/M Phase |  |
| --- | --- | --- | --- | --- | --- | --- |
|  | F | p-value | F | p-value | F | p-value |
| Group<br>(0=Control, 1=Case) | 0.002 | 0.963 | 0.003 | 0.958 | 6.988 | 0.0171 |
| Fluoxetine Treatment<br>(0=Untreated; 1=Treated) | 2.511 | 0.131 | 0.078 | 0.783 | 5.967 | 0.0258 |
| Group*Fluoxetine<br>Treatment Interaction | 0.679 | 0.421 | 1.623 | 0.22 | 1.272 | 0.2751 |

**Table 2:** Post Hoc Test (Tukey’s HSD) for the Percentage of cells in G2/M Phase.

| Pairwise Contrast | Mean<br>Difference | 95% Confidence<br>Intervals |  | Adjusted<br>P-Value |
| --- | --- | --- | --- | --- |
|  |  | Lower | Upper |  |
| Treated:Case - Untreated:Case | -1.315 | -4.400 | 1.769 | 0.661 |
| Untreated:Control - Untreated:Case | 3.288 | -0.073 | 6.650 | 0.057 |
| Treated:Control - Untreated:Case | -0.055 | -3.417 | 3.306 | 1.000 |
| Untreated:Control - Treated:Case | 4.604 | 1.242 | 7.965 | 0.004 |
| Treated:Control - Treated:Case | 1.260 | -2.102 | 4.621 | 0.743 |
| Treated:Control - Untreated:Control | -3.344 | -6.961 | 0.273 | 0.078 |

### 3.2 Cell cycle from SRI-nonresponders showed decreased proliferative capacity

Comparing among OCD cases, we found LCLs from clinical non-responders to SRI treatment had a significantly lower percentage of cells in the G2/M phase compared to treatment responders (10.131.56 vs 13.781.70, t=-3.088, p=0.023). No significant difference was found in the percentage of cells in other phases of the cell cycle, either between the groups or within groups, with or without *in vitro* fluoxetine (Table 1, Figure 1).

**Figure 1.**
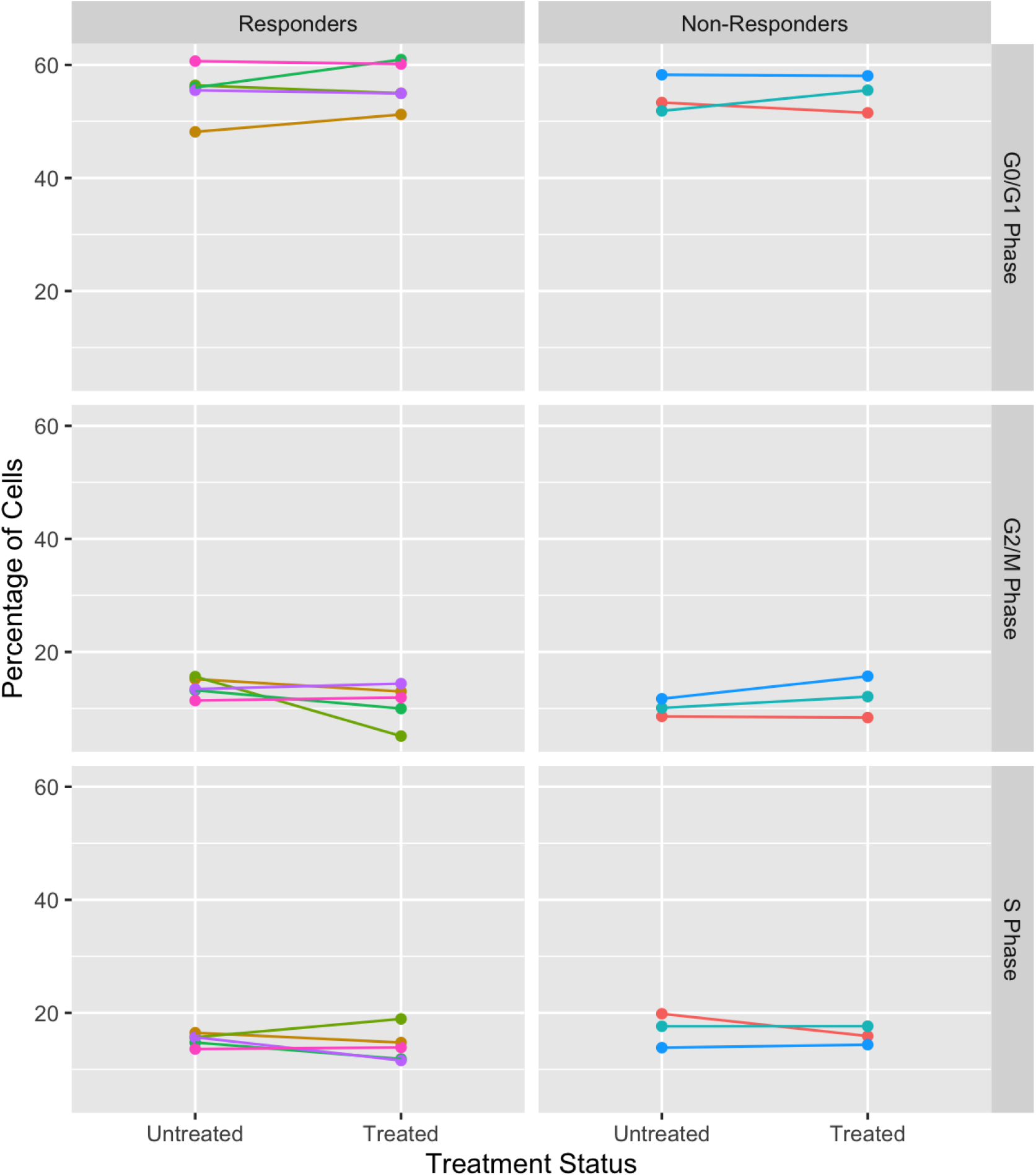
Difference between clinical treatment responders (n=5) and non-responders (n=3) in the percentage of LCLs in each phase of the cell cycle, with and without fluoxetine treatment.

Previous studies have shown fluoxetine reduces the number of cells in the G2/M and S phase (Lin et al., 2016; Solek et al., 2019). These effects were evident at 24 hrs with 10μM fluoxetine, which was the reason for choosing these parameters for the present study. Following *in vitro* fluoxetine treatment (10μM for 24 hrs), we observed an overall reduction in the proportion of cells in the G2/M phase in cases and controls LCLs. The *in vitro* effects of fluoxetine need further exploration with a longer duration of treatment and larger sample numbers.

## 4. Discussion

A critical and yet unanswered question in Psychiatry is how might clinical phenotypes manifest in terms of cellular readouts. Such objective biomarkers will help bring about a better understanding of psychiatric disorders, which thus far are principally known in terms of clinical signs and symptoms. The present study was undertaken with this overarching aim as our guiding principle.

The primary finding in this study is that there is a reduction in the number of proliferative LCLs from OCD cases. Since the number of cells in other phases of cell cycle is the same across experimental paradigms, the reduction in G2/M phases could indicate cell cycle arrest. Physiologically, dividing cells undergo cell cycle arrest at this stage if DNA damage is detected. Though we did not probe for DNA damage in these LCLs, interestingly, Alici et al. (2016) did demonstrate oxidative DNA damage in peripheral blood in OCD patients.

An obvious next question is how might a finding in peripheral blood, or cell type derived from peripheral blood (LCL) be representative of mechanisms related to a brain disorder. Considering that brain biopsies from patients are not a realistic option, lymphocytes are a convenient model system in which to probe cellular functions. Substantial literature already exists to corroborate this principle for bipolar disorder (reviewed in Viswanath et al. 2015); though comparable evidence is largely wanting for OCD. Given that there are overlaps in cellular mechanisms related to severe mental illnesses, we wanted to see if LCLs derived from OCD patients display abnormal cellular phenotype. Disturbances in cell cycle, as we report here in OCD, have earlier also been reported for depression (Vaidya et al. 2007), schizophrenia (Wang et al. 2010, Ashok et al., 2019) and bipolar disorder (Paul et al., 2020). Collectively, this suggests that cell cycle dysregulation is a common molecular phenotype across clinically diverse psychiatric disorders.

## Data Availability

Available by contacting the corresponding author

## Acknowledgements

This work was supported by “Accelerator program for discovery in brain disorders using stem cells” (BT/PR17316/MED/31/326/2015) (ADBS) from Department of Biotechnology (DBT) and Pratiksha Trust; and SCIENCE & ENGINEERING RESEARCH BOARD (SERB) project “Dissecting the biology of lithium response in human induced pluripotent stem cell derived neurons from patients with bipolar affective disorder” (FILE NO. ECR/2016/002076).

